# Flexible-circuit-based 3-D aware modular optical brain imaging system for high-density measurements in natural settings

**DOI:** 10.1101/2024.03.01.24302838

**Authors:** Edward Xu, Morris Vanegas, Miguel Mireles, Artem Dementyev, Meryem Yücel, Stefan Carp, Qianqian Fang

## Abstract

**Significance:** Functional near-infrared spectroscopy (fNIRS) presents an opportunity to study human brains in every-day activities and environments. However, achieving robust measurements under such dynamic condition remains a significant challenge.

**Aim:** The modular optical brain imaging (MOBI) system is designed to enhance optode-to-scalp coupling and provide real-time probe 3-D shape estimation to improve the use of fNIRS in everyday conditions.

**Approach:** The MOBI system utilizes a bendable and lightweight modular circuit-board design to enhance probe conformity to head surfaces and comfort for long-term wearability. Combined with automatic module connection recognition, the built-in orientation sensors on each module can be used to estimate optode 3-D positions in real-time to enable advanced tomographic data analysis and motion tracking.

**Results:** Optical characterization of the MOBI detector reports a noise equivalence power (NEP) of 8.9 and 7.3 pW/ Hz at 735 nm and 850 nm, respectively, with a dynamic range of 88 dB. The 3-D optode shape acquisition yields an average error of 4.2 mm across 25 optodes in a phantom test compared to positions acquired from a digitizer. Results for initial *in vivo* validations, including a cuff occlusion and a finger-tapping test, are also provided.

**Conclusions:** To the best of our knowledge, the MOBI system is the first modular fNIRS system featuring fully flexible circuit boards. The self-organizing module sensor network and automatic 3-D optode position acquisition, combined with lightweight modules (18 g/module) and ergonomic designs, would greatly aid emerging explorations of brain function in naturalistic settings.

## 1 Introduction

The widespread adoption of functional neuroimaging techniques has fundamentally advanced our understanding of how the human brain perceives, interprets, and responds to stimuli.^1^ However, brain activation exhibits complex patterns and dynamics, many of which are only apparent when studied in their native environments.^2^ This has posed a challenge for many established neuromonitoring techniques, such as functional magnetic resonance imaging (fMRI)^3^ and magnetoen-cephalography (MEG),^4^ which, despite tremendous technological innovation over recent years, still suffer from a key limitation – the need for subjects to remain relatively motionless within a highly confined space. Such limitations have greatly restricted our ability to understand the human brain in everyday situations and interactions.^5, 6^ Although another commonly used neuroimaging technique, electroencephalography (EEG), complements fMRI/MEG with excellent portability, it is limited by relatively poor spatial resolution.^5, 7^ These inherent disadvantages of gold-standard neuromonitoring modalities have motivated active exploration and development of new techniques that are more suited to measuring brain activities in naturalistic and dynamic environments.

In recent years, researchers have turned to functional near-infrared spectroscopy (fNIRS) to address this technological gap.^8^ FNIRS is an emerging functional neuroimaging technique that measures the hemodynamic changes associated with brain activity^9^ using safe, non-ionizing nearinfrared light.^8^ With its relatively low cost and ability to probe a rich set of physiological parameters, fNIRS has seen rapid adoption in recent years^6, 10^ and has been used in studies ranging from psychiatric conditions,^11, 12^ language,^13–15^ cognitive neurodevelopment,^16–19^ stroke recovery,^20^ education,^21^ pain detection,^22^ and even brain-computer interfaces.^23–25^ However, many early fNIRS studies employed mobile, cart-sized instruments that housed the source and detector units and optical fibers that transmit light to and from the a subject’s scalp.^26–29^ Although such fiber-based fNIRS systems are significantly more portable than conventional fMRI and MEG, the fragility of optical fibers and physical constraints upon subject mobility still preclude them from investigating brain activity in unrestricted environments.^1^

In response to this, fNIRS devices featuring improved wearability and portability have been actively developed over the past decade and deployed in both research and clinical settings.^30^ In particular, the rapid rise of modular fNIRS architectures,^31, 32^ alongside a diverse range of lightweight and wearable fNIRS instruments,^33–39^ represents a promising step towards extending fNIRS-based brain monitoring to address the aforementioned challenges. Modular fNIRS systems are wearable probes comprised of repeating source and detector circuits called modules.^40^ The use of repeating modules not only greatly simplifies system fabrication and lowers overall production costs, but also offers improved flexibility in probe design to readily adapt to the diverse coverage requirements and regions-of-interest (ROIs) of different fNIRS experiments.^41^ In addition, wearable fNIRS devices have increasingly replaced cumbersome fiber optics with compact sources and detectors placed at the scalp surface to allow for greater optode density and more robust system designs. Modular fNIRS architectures with the ability to measure from both intra- and inter-module (optodes on different modules) source-detector (SD) pairs further improve spatial sampling density and overlapping channels, which have been shown to enhance both spatial resolution and depth specificity.^38, 42^ However, compared to traditional “monolithic” fNIRS systems, modular headgear require additional considerations, including optode layout design and modular connectivity, to optimize measurement coverage. Our group has recently released an open-source toolbox specifically addressing this need.^43^

While the transition towards modular and wearable fNIRS systems represents an important initial step towards achieving robust measurements of brain activity in naturalistic settings, additional challenges remain to be addressed. For example, the increased subject and headgear movement under dynamic experimental conditions can result in strong motion-related artifacts.^5, 44^ While some of these artifacts can be detected and corrected using signal processing techniques^45, 46^ or alleviated through mechanical approaches like glue^47^ or spring-loaded optode designs,^48^ effectively and efficiently coupling sources and detectors to the head remains a key obstacle.^49^ Additionally, model-based tomographic image reconstruction techniques using data from multiple channels overlapping the same area have been shown to improve the spatial localization accuracy, signal contrast, and measurement robustness of recovered brain activations.^50, 51^ However, this approach requires 3-D optode positions^52^ at the very least and, in some cases, 3-D anatomical scans. The typical approach for acquiring 3-D optode positions involves manual measurements using 3-D digitization systems, providing a snapshot of optode locations without accounting for movement or positioning changes over the course of an experiment.^37, 53^ More recent solutions leveraging photogrammetry offer greater portability and constant sampling over time, but still require a set of external cameras that restrict mobility to areas within the cameras’ field of view.^38, 52^ Integrating automated, real-time measurements of 3-D optode positions into a wearable fNIRS system would further assist motion correction and advance tomographic fNIRS to main-stream use.

To address the aforementioned challenges, we report here a lightweight, re-configurable, highdensity fNIRS modular optical brain imager (MOBI) system using head-conforming flexible/bendable circuits with integrated 3-D optode position tracking capabilities.^54^ This ultra-compact and fiberless system adopts a diamond-shaped flexible printed circuit board (fPCB) module that improves the robustness of optode-scalp coupling.^55, 56^ Additionally, a dense peer-to-peer (P2P) network automatically determines module-to-module connectivity and topology, facilitating the implementation of spatial multiplexing strategies that increase data acquisition frame rates. Finally, each flexible MOBI module contains an inertial measurement unit (IMU), which can be used in combination with the automatically determined connection topology to produce real-time, 3-D tracking of optode positions. This built-in position-sensing capability greatly simplifies tomographic fNIRS data acquisition, shortens setup times, and provide real-time metrics for assessing optode motion.

In the subsequent sections, we provide an overview of the MOBI system, including technical specifications for the MOBI modules and supporting hardware components as well as implementation strategies for novel features such as connection topology recognition, IMU-based optode positioning, and data acquisition speed improvement through spatial multiplexing. We further characterize the head-conformability of flexible-versus rigid-circuit probes, assess the accuracy of automatically determined optode positions, and provide preliminary validation of system performance in phantom and *in vivo* experiments.

## 2 Methods

### 2.1 Module design

A MOBI module consists of a double-sided fPCB fabricated on polyimide-based films [Fig. 1(c)], with each module housing two detectors (OPT101, Texas Instruments, USA) with built-in transimpedance amplifiers and 3 dual-wavelength light-emitting diode (LED) sources at 735 and 850 nm (Marubeni, Japan). The LEDs are driven by a single programmable constant current driver (LT3092, Analog Devices Inc., USA) capable of delivering 0-100 mA of current coupled to a digital-to-analog converter (DAC, Microchip Technology Inc, USA) and controlled by a digital multiplexer (NX3L4051, NXP Semiconductors, Netherlands) using a spatial multiplexing encoding strategy as detailed in Section 2.4.2 (Fig. 2).

**Fig 1.**
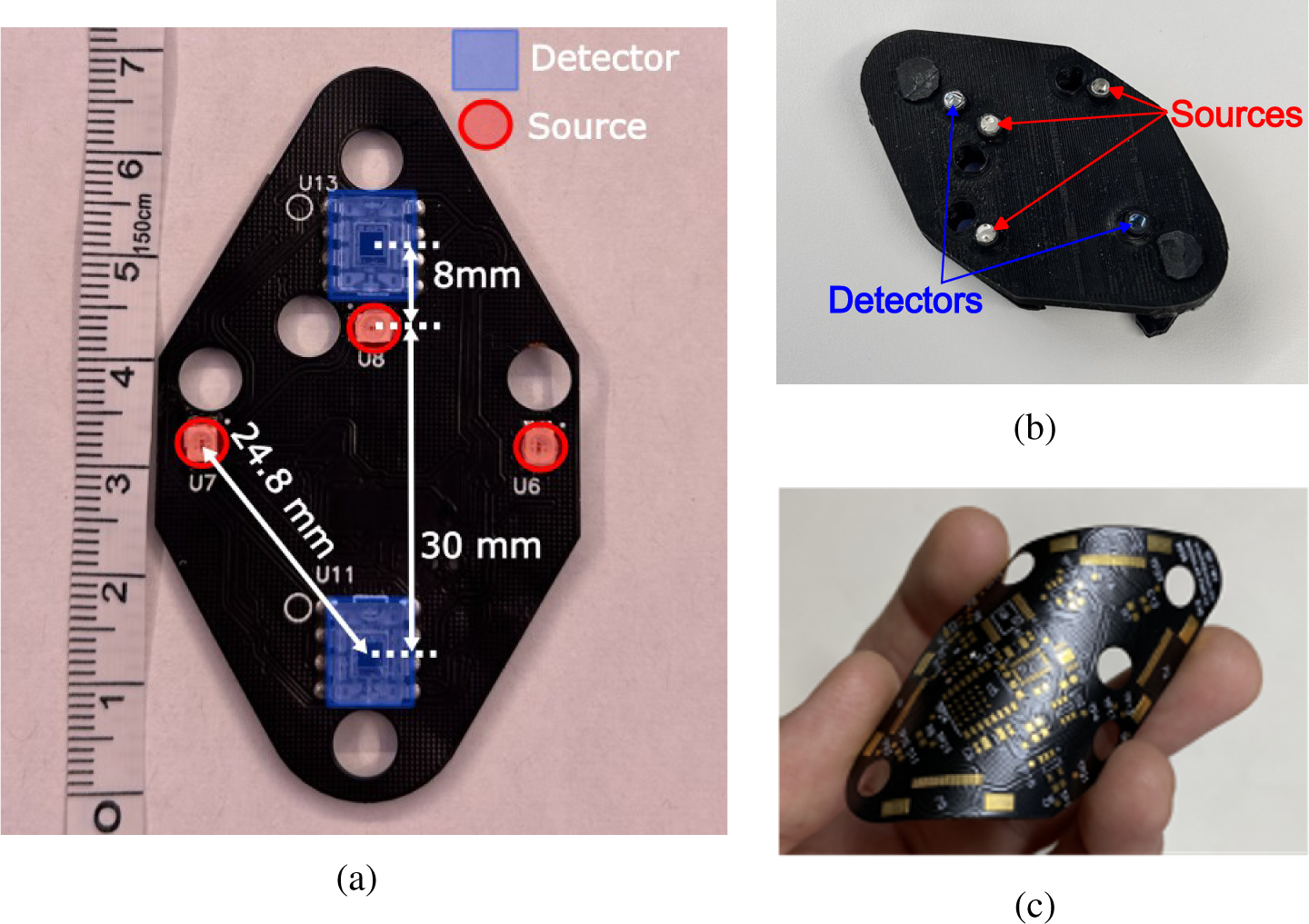
Photos of the MOBI module showing the (a) top view without a silicone cover, (b) bottom view showing light guides attached to optodes and a black silicone enclosure, and (c) MOBI flexible-circuit-board before adding components.

**Fig 2.**
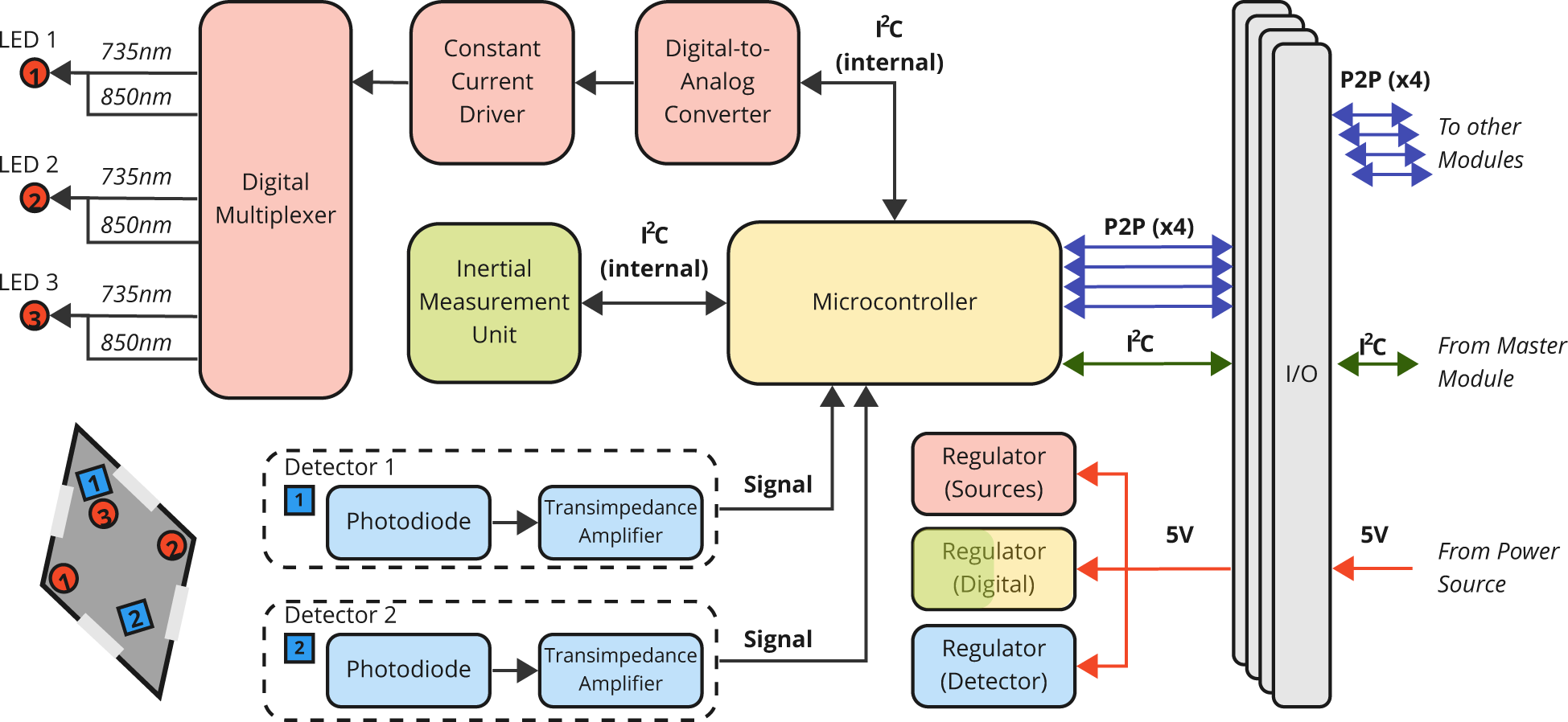
Schematic diagram of a single MOBI module. The microcontroller uses an internal inter-integrated circuit (I^2^C) protocol to communicate with components on a single board. A peer-to-peer (P2P) network allows communication between neighboring modules. Associated components are color-coded: red for sources, blue for detectors, green for the IMU, and yellow for the microcontroller.

The positions of the LEDs and detectors are labeled on Fig. 1(a). This arrangement produces one 8 mm, one 30 mm, and four 24.5 mm dual-wavelength channels within a single module (referred to as intra-module channels hereafter). In addition, a 3 mm diameter acrylic optical fiber (Edmund Optics, USA), is attached to each source and detector, serving as a light guide to couple light to and from the scalp [Fig. 1(b)]. At the center of each module, we also mount a 9-axis IMU that tracks the module’s orientation (Bosch Sensortec, Germany). Additionally, three low-dropout (LDO) voltage regulators are used to control the power source voltage for the source, detector, and auxiliary measurement components. All optodes and sensors on the MOBI module are controlled by an onboard system-on-a-chip (BC832, Fanstel Corp, USA) with an integrated microcontroller (nRF52832, Nordic Semiconductor, Norway). The microcontroller also incorporates a 12-bit, 2-channel analog-to-digital converter (ADC) with 8 built-in gain settings, which change the effective input range of the ADC, that samples from the detectors and IMU. Each optical measurement averages 175 samples, providing a single-channel, single-wavelength acquisition time of 7.5 ms and a modular frame rate, including all six dual-wavelength channels, dark measurements, and IMU measurements, of 11 Hz. An inter-integrated circuit (I^2^C) protocol is used to communicate between various components (Fig. 2).

The five optodes (3x LEDs and 2x detectors) are located on the side of the module facing the scalp while all other electronic components are placed on the opposite side to assist with signal isolation, heat dissipation, and comfort. MOBI modules have a channel density of 0.56 dual-wavelength channels per square centimeter. We designed our module using a diamond or dual-equilateral-triangle shape with an edge length of 5 cm, a choice motivated by the superior tesselation of curved head surfaces with triangular meshes. Each module is encapsulated in a cast flexible black silicone enclosure to provide a smooth touch, blockage of stray light, and protection for electronic components during use. Each module has four flexible printed circuit (FPC) connectors that allow multiple modules to be connected using FPC cables [Fig. 1(a)]. A peer-to-peer (P2P) serial network protocol allows each module to communicate with up to four connected neighboring modules. Five 8 mm diameter holes next to each optode are used for securing modules to a headgear and potentially hair adjustment. Each module, including all components, light guides, and the silicone cover, has a total weight of around 18 grams.

### 2.2 System architecture

A compact master node, measuring 45*×*30 mm^2^, is used to connect to and control an arbitrary number of daisy-chained MOBI modules (Fig. 3). When the master node is powered by a laptop using a single universal serial bus (USB) cable, it can support up to 6x MOBI modules. An external power supply can be used instead when using more modules is desired. In addition, the master node utilizes I^2^C communication to supply power to and acquire data from each module [Fig. 3(a)]. It incorporates a microcontroller development board (Teensy 4.0, PJRC, USA), a voltage regulator, an FPC connector, a 2-pin Japan Solderless Terminal (JST) connector, and two switches. A micro-USB cable connects the master board to a computer for serial communication. The switches are used to manually toggle the power supply (between micro-USB and an external battery) to the modules and the JST connector allows an external trigger board [Fig. 3(c)] to be connected for synchronizing auxiliary signals such as experimental triggers. This trigger board is based on a simple microcontroller (ATmega328P, Microchip, USA) and transmits transistor-transistor logic (TTL) signals to the master node. The master node and trigger boards are individually encased inside 3-D printed covers.

**Fig 3.**
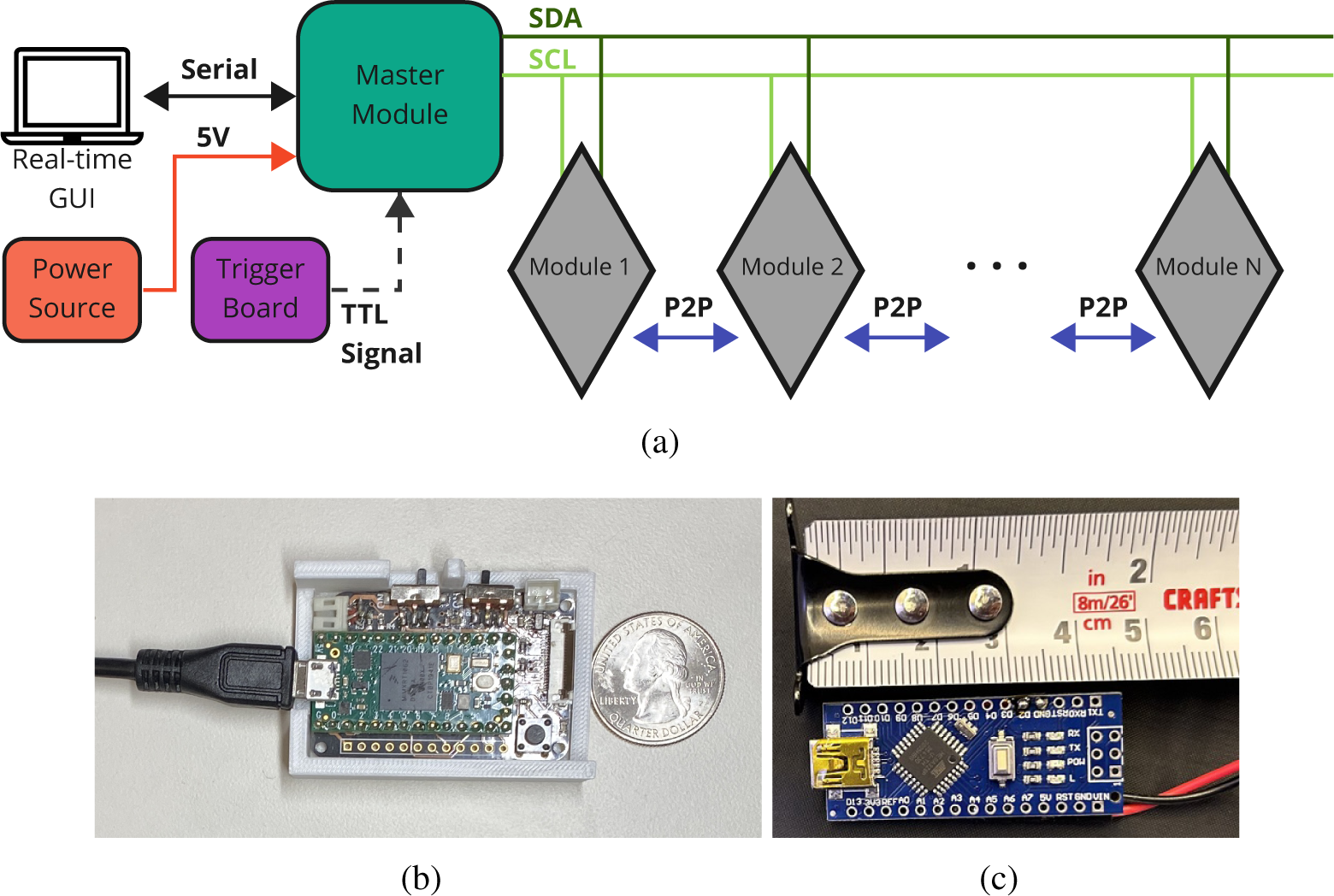
Schematic diagram of (a) a multi-module MOBI probe. Optional external power source and trigger board not shown. We also include in (b) a photo for the master node with its circuit board exposed and (c) a photo of the hardware trigger board.

Data acquisition of a MOBI probe is controlled through a MATLAB (Mathworks, Natick, MA) based graphical user interface (GUI) on the computer, which allows users to perform pre-experimental calibration and setup, acquire and visualize measurements, and save data into the community-driven Shared Near Infrared Spectroscopy Format (SNIRF).^57^ Optical signal quality can be manually adjusted from the GUI by setting the source current and detector gain settings (ranging from 1/6 to 4*×* gain), with real-time signal display providing instantaneous feedback including visual indicators for channels near saturation or noise floor. In addition, the GUI accepts software triggers transmitted through LabStreamingLayer (LSL),^58^ a middleware for synchronizing experimental data streams, and provides visual cues of trigger arrival within the data visualization windows. MOBI also supports wireless communication of data and user-inputted commands between the master node and control computer through a Wi-Fi co-processcor breakout board (ESP32, Adafruit Industries, USA) for the master node and transmission control protocol/internet protocol (TCP/IP) communication. During Wi-Fi based data acquisition, a MOBI probe can be entirely powered by a battery without needing to physically connect to a computer.

### 2.3 Automatic features

#### 2.3.1 Automatic connection topology recognition

Our MOBI system has the ability to automatically recognize the connection topology between all modules to determine the number of modules and the orientation of each module in an arbitrary probe. An animation demonstrating this feature is shown in Fig. 4. Modules can be dynamically added to an existing probe by connecting one of its four FPC connections with another module’s FPC connector using an FPC cable. Upon start-up, each module samples all four of its P2P communication channels to determine if one or multiple modules are connected to its FPC connectors. This sampling of inputs and outputs allows the P2P serial network to automatically determine how each module is positioned relative to others. Each module is also programmed with a global module ID. With this information, the computer can determine the spatial orientation of each module in the probe based on the connection topology and the master node can implement spatial multiplexing patterns generated using the MOCA toolbox.^43^

**Fig 4.**
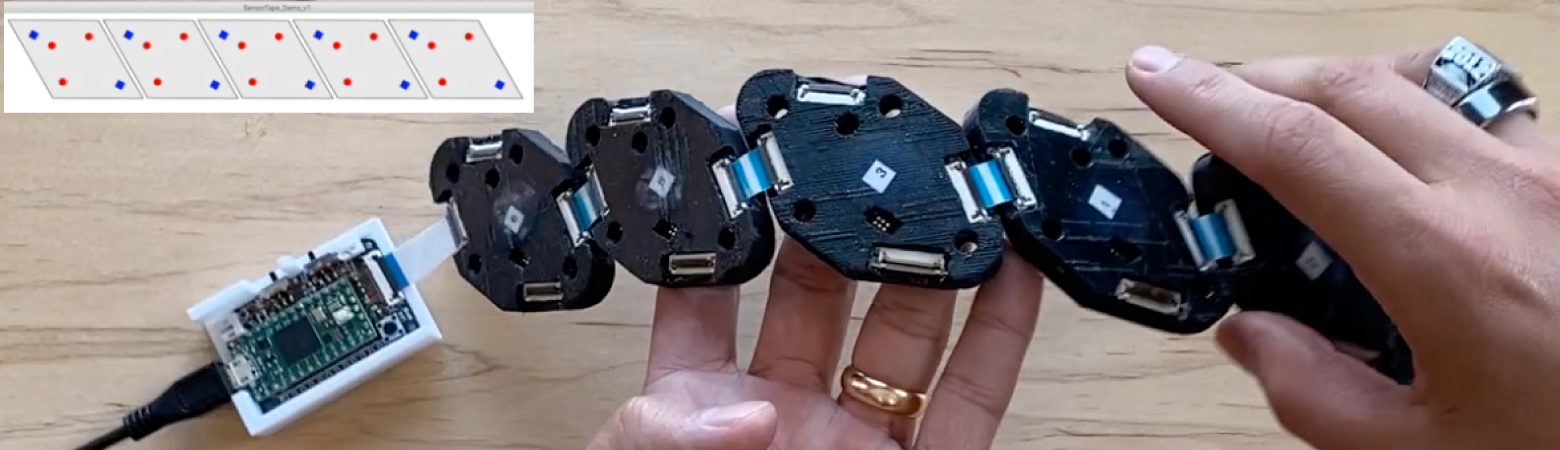
Demonstration of automatic determination of module-to-module connectivity, shown in the top-left overlay, using two five-module configurations. [Video 1 (MP4, 5 MB)]

#### 2.3.2 Automatic 3-D optode position estimation using IMU

Additionally, our MOBI system can automatically determine the 3-D position of each source and detector in the probe without the use of an external digitizer. A demonstration of this feature is also provided in Fig. 4. The previously determined connection topology, known FPC cable lengths, and orientation measurements acquired from each onboard IMU are used together to generate a 3-D piece-wise spherical model to estimate the surface upon which the probe is placed. The detailed methods are described in Section 2.4.4. Module geometry information and optode layouts within a module can then be superimposed upon this surface to derive the 3-D location of each optode. IMU measurements are also recorded over the duration of an experiment and can be used for robust temporal signal rejection through real-time monitoring of optode movements during use.

#### 2.3.3 Spatial multiplexing groupings

Our MOBI modules utilize a spatial multiplexing encoding strategy. Source LEDs are grouped into spatial multiplexing group (SMG) based on their spatial distributions during the probe design phase using our MOCA toolbox.^43^ LEDs within the same SMG can be illuminated simultaneously due to sufficient spatial separation, provided by the user when creating the probe, and negligible cross-talk while LEDs across SMGs are illuminated sequentially. This approach allows us to accelerate data acquisition by a factor referred to as spatial multiplexing ratio, or SMR, defined as the totally number of source LEDs in a probe divided by the number of SMGs. For example, an SMR of 2 suggests that the data acquisition is 2-fold faster than turning on every source sequentially.

### 2.4 System characterization protocols

#### 2.4.1 Optical performance characterization

The MOBI system’s signal quality is evaluated using several performance metrics. Detector sensitivity is characterized using a dual-axis filter wheel setup, shining light from a source LED through 36 levels of attenuation ranging from neutral-density filter with optical density (OD) ratings from 0 to 4.8 and sampling with a MOBI detector. From these measurements, we can calculate noise equivalence power (NEP), defined as the value at which the optical signal and noise are of equal magnitude. This also allows us to estimate the detector’s dynamic range as the ratio of maximal detectable power over the minimal detectable signal (i.e. NEP). Additionally, we also characterize MOBI’s performance on the probe-level by measuring the signal-to-noise ratio (SNR) at various source-detector separations (SDSs). A linear 3-module probe was created, providing SDSs ranging between 24 - 85 mm. Measurements were performed on an optical phantom (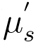 = 4.7 cm*^−^*^1^ and µ*_a_* = 0.063 cm*^−^*^1^ at 830 nm), acquiring 100 samples for each channel and wavelength to compute the respective SNRs. Both the source LED current and detector gain settings are tuned to maximize the number of usable channels.

#### 2.4.2 Sampling rate enhancement using spatial multiplexing

The ability to improve probe sampling rates utilizing spatially multiplexed groups (SMG) is conceptually demonstrated via our MOCA toolbox using three distinct probe configurations as examples. All tested configurations contain identical numbers of modules (N = 5) and only differ in their spatial connections. Using MOCA, we calculate the number of valid intra- and inter-module channels that fall within an SDS threshold, set to 35 mm for this analysis, and determines the lowest number of SMGs needed. We compare the channel numbers, SMGs and probe data frame rate from these 5-module probes to exemplify the importance of spatial multiplexing in modular probe design.

#### 2.4.3 Optode-scalp conformity improvement

We previously fabricated rigid PCB versions of our flexible modules with the same dimensions and diamond shape, providing an opportunity to quantitatively assess the enhancement in optode-scalp conformity within the constraints of our module design. We first 3-D print a hemisphere phantom of radius 100 mm to simulate a simplified head-surface. Identical V-shaped 5-module probes are then created using either rigid and flexible modules and placed atop the hemisphere and held in place using a mesh hair net. The 3-D positions of all 25 optodes are manually digitized for both rigid and flexible probes using a 3-D digitizer (Fastrak, Polhemus, Colchester, VT) and then used to compute the distance of each optode to the center of the sphere. Four landmarks along the bottom circumference of the hemisphere are also digitized to estimate the position of the sphere center. The distances between the digitized optode positions from either probes are compared to the expected radial distance (105 mm, consisting of the sphere radius and the module thickness) when the modules are perfectly conforming to the surface.

#### 2.4.4 Automated optode localization algorithm and accuracy assessment

A fast algorithm was developed to recover 3-D optode/probe positions using the acquired IMU readings and the known probe connection topology, explained in a graphical illustration shown in Fig. 8(a). The 3-D optode positions are estimated in two steps. First, we select any of the modules, preferably the middle module of a series of inter-connected modules, as the reference (R) with a specified origin (p*_R_*), and estimate the center position of the module (P) immediately adjacent to the reference module using the below algorithm: we first use the quaternion readings from the IMUs in modules R and P to compute the angle of rotation between them, θ, and their respective normal vectors, (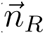) and (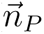), which are the quaternion rotation applied to the (0, 0, 1) unit vector. By assuming that the surface between every adjacent module can be approximated by a spherical surface, we can use θ and the known cable length (L*_P,R_*) between the two modules (55 mm in this experiment) to determine the radius of the sphere, r = L*_P,R_*/θ. The module center of P can be then computed by

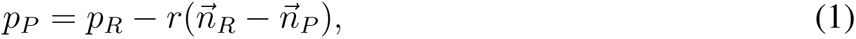

where, p*_P_* and p*_R_* are the respective module center positions; 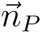 and 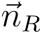 are the respective normal vectors; r is the radius of the sphere. Once we determine the center positions of P using R as reference, we continue applying the above algorithm to all subsequently connected modules and propagate the positions to every connected modules. To enhance the accuracy of our estimation, we constrain modules that are connected in a straight line topologically to the same plane by averaging the pair-wised planes between each estimated module and the reference module and projecting the quaternion-derived normal vectors onto this plane. Using the updated normal vectors, we recompute the module center positions starting again from the reference module.

In the second step, we estimate the 3-D positions of the optodes from the center of each module based upon the 2-D module layout and radii used to estimate module positions in the previous step. This is achieved by using the module’s local coordinate system, with the origin located at the module center with the short-axis serving as the x-axis. The 2-D distance between each optode to the module center is known, defining the arc length between the origin and the optode over a local spherical surface with a radius calculated as the average of the pair-wised spherical radii estimated in the previous step. Once the 3-D positions are computed in the local coordinate system, they are then translated to the probe’s world-coordinate using the module center’s position and normal vector.

The accuracy of IMU-derived 3-D optode positions is assessed using the same flexible V-shaped 5-module probe and hemisphere phantom used in the previous section. Manual digitization of optode positions are again performed using the Polhemus digitizer and repeated five times to produce the “ground-truth” from the averaged positions and the standard error of multiple digitizations from a single operator. The central module lying at the tip of the V-shaped probe serves as the reference module for IMU-derived optode locations and for registering IMU-derived and manually digitized positions.

#### 2.4.5 *Initial* in vivo *validation*

The MOBI system’s ability to measure hemodynamic changes is evaluated in human subjects using two experimental protocols. First, we perform a blood pressure cuff occlusion experiment on a adult male subject of 30-35 years of age while collecting simultaneous measurements using MOBI and a commercial fNIRS system (Brite23, Artinis) as a reference to assess the trend and magnitude of recovered physiological changes. A single MOBI module and single channel Artinis probe are placed on the medial forearm. A blood pressure cuff is positioned on the upper arm, which is rested at the same level as the heart. The cuff is first inflated to 100 mmHg and then to 220 mmHg for 75 seconds each to achieve venous and arterial occlusions, respectively. Measurements from the 30 mm channels of both the MOBI and Artinis systems are acquired simultaneously at 10 Hz in a dark room.

In addition, the evoked response for a finger-tapping experiment is measured in a right-handed adult male of 30-35 years of age. The task consists of ten repetitions of 20-second task periods, during which the non-dominant hand sequentially taps the thumb to the index, middle, ring, and little fingers of the same hand, and 25-second rest periods, with 60-second rest periods added before and after the entire task. The subject sits in a comfortable position with their eyes closed during the entire experiment and is verbally instructed to start and stop tapping. During rest, the subject is instructed to place both hands on their laps. A probe consisting of two MOBI modules is created and symmetrically placed over the ipsilateral and contralateral primary motor cortices,^59^ positioned halfway between C1 and C3 for the left hemisphere and between C2 and C4 for the right hemisphere using the EEG 10-20 system.^60^ The modules are mounted and secured using a flexible, 3-D printed head-cap developed in-house [Fig. 10(a)].^61^ Our cap contains embedded 10-10 position markers computed from an adult atlas to guide module placement and features a wire-frame mesh to securely and conveniently anchor MOBI modules in any position using 3-D printed locking pins and clips. This combination of wire-frame and locking pins also allows for the rapid re-configuration of MOBI modules between experimental probe designs. A black light-blocking fabric is placed over the head-cap to further block ambient light.

Acquired data are converted into the SNIRF format and loaded into Homer3^62^ for processing. A simple processing stream is employed – the raw data is converted to optical density (OD), a band-pass filter with pass-band frequencies between 0.01 and 0.4 Hz is applied, and the OD is converted to blood concentration using the modified Beer-Lambert law with the partial path-length factor set to 1.^63^ The hemodynamic response function (HRF) is then computed with the general linear model (GLM) using the short separation most closely correlated with each measurement channel to regress physiological signals.^64^

## 3 Results

### 3.1 System characterization

Characterizing the optical sensitivity of the MOBI modules, shown in Fig. 5(a), demonstrates a noise equivalent power of 8.90 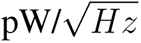 at 735 nm and 7.28 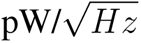 at 850 nm. In addition, the detector saturates at 250 nW and 220 nW for 735 and 850 nm, respectively, providing a dynamic range of around 88 dB for both wavelengths. Fig. 5(b) shows the SNR versus source-detector separation over an optical phantom. For this measurement, the source current is attenuated to 50% and no detector gain is applied to maximize the number of non-saturated channels. Under these conditions, intra-module channels (24.8 mm and 30 mm) exhibit an SNR of 65-72 dB and SNRs of greater than 40 dB are measured in source-detector separations of up to 50 mm.

**Fig 5.**
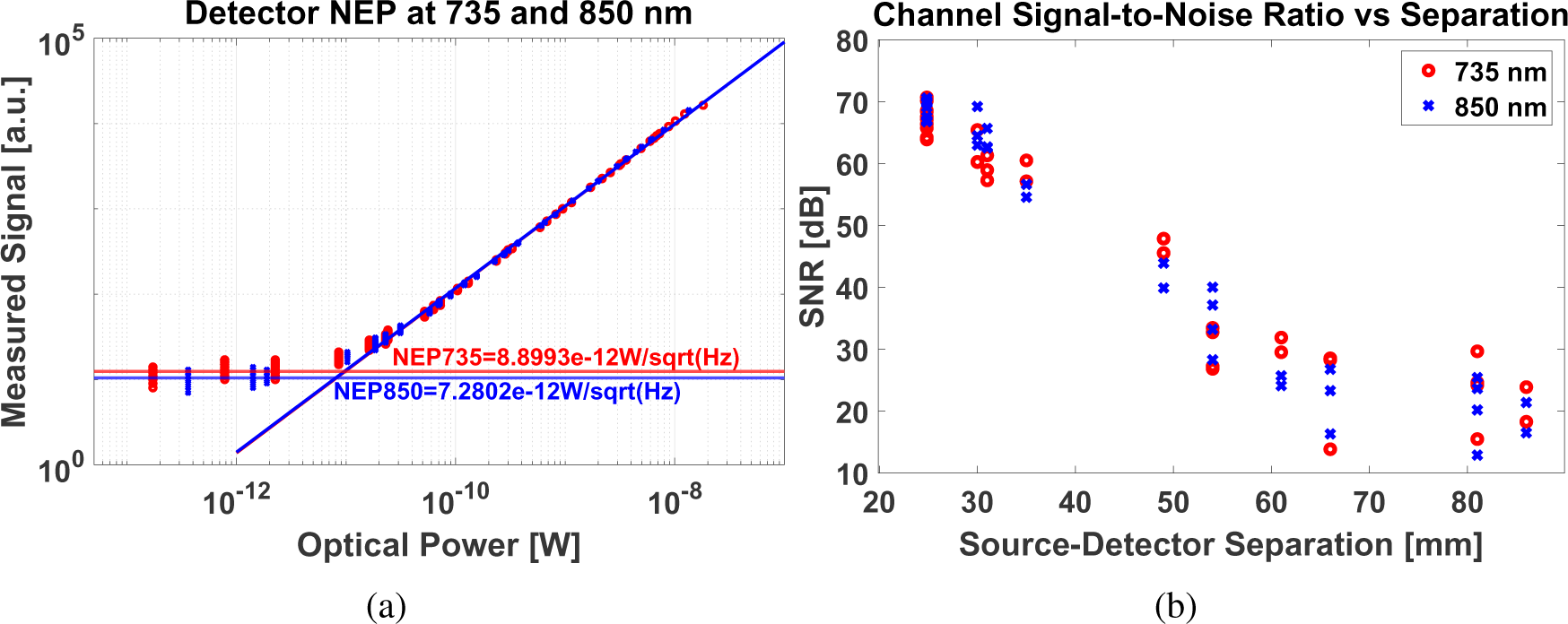
System characterization showing (a) measured signal vs. incident optical power, providing noise equivalent power at both 735 and 850 nm and (b) signal-to-noise ratio of versus source-detector separation from a 3-module probe, with each point representing a unique channel.

Although we do not have enough completed modules to physically create and test a full-head MOBI probe, we use our MOCA toolbox and MOBI module specifications to predict some of the key metrics for a full-head probe configuration. Based on the head surface area computed from an adult atlas, we estimate that a total of 20 MOBI modules is needed. Using the MOCA toolbox, this full-head probe provides 372 dual-wavelength channels including both intra- and inter-module channels. This increases the achievable channel density to 1.72 channels per square centimeter. Additionally, with MOBI’s spatial multiplexing encoding strategy and an SDS cutoff of 50 mm, this full-head probe results in 14 spatial multiplexing groups and a full frame rate of 2.4 Hz. In terms of power, assuming each source is operated at the maximum 100 mA to produce an illumination of 12.3 and 9.6 mW at 735 and 850 nm, respectively, at the output of the coupler,^65^ the total power draw of the full-head probe is estimated at 2.31 W, resulting in a 2.85-hour battery life when a 3.7 V 2,000 milliampere-hour (mAh) battery is used.

### 3.2 Spatial multiplexing groups and data acquisition speed

As described in Section 2.4.2, three example 5-module probe layouts are tested and shown in Fig. 6. The predicted performance using MOCA for each probe is compared in Table 1. When using a sequential acquisition scheme, the full frame-rate of the probe is inversely proportional to the number of sources in the probe, resulting in 2.2 Hz for all 3 configurations. Using MOCA, we can divide the sources into SMGs so that LEDs belonging to different SMGs can be turned on simultaneously. In Table 1), we report different probe-level metrics reported by MOCA. It is evident that SMGs can significantly enhance the full frame rate of probes while module layouts that are more spatially packed require higher numbers of SMGs.

**Fig 6.**
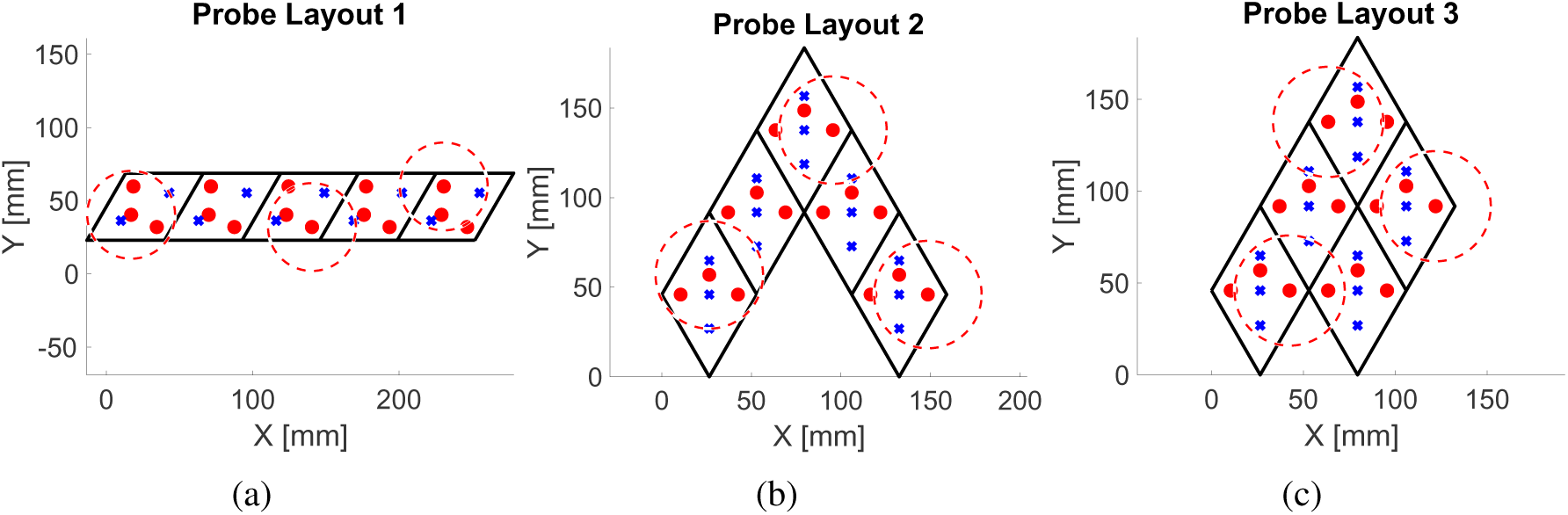
Three example probe layouts composed of five identical MOBI modules. Optodes are represented by small red circles (sources) and blue crosses (detectors). Each layout has multiple spatial multiplexing groups determined based on the global proximity of sources to each other. Red dashed circles show which sources are simultaneously on for each layout’s first spatial multiplexing group.

**Table 1.** The full frame rate of a probe depends on the layout of the modules within a probe. The three layouts in Fig. 6 result in the channels, groupings, and full frame rates below.

|  | Layout 1 | Layout 2 | Layout 3 |
| --- | --- | --- | --- |
| Number of modules | 5 | 5 | 5 |
| Frame rate for sequential acquisition [Hz] | 2.2 | 2.2 | 2.2 |
| Number of channels | 38 | 53 | 55 |
| Number of spatial multiplexing groups | 5 | 7 | 7 |
| Improvement ratio | 3.0 | 2.2 | 2.2 |
| Spatial encoding full frame rate [Hz] | 6.6 | 4.8 | 4.8 |

### 3.3 Quantifying optode conformity to head surface

Fig. 8(b) shows a photo of the five-module probe, constructed using either rigid or flexible modules (see Figs. 7a-b), pressed upon a hemispherical phantom with a mesh hair-net. If the module is perfectly conforming to the phantom surface, the distance between the top of the module and the center of the sphere is expected to be 105 mm, consisting of the 100 mm dome radius and 5 mm thickness of the module. For the rigid-circuit probe, the average measured distance is 107.0±3.7 mm, with nine optodes exhibiting errors of more than 5 mm from the expected distance. The use of flexible probes lowers the average measured distance to 104.8±2.8 mm. In addition, measured distances for the flexible modules are more centrally distributed [Fig. 7(f)] when compared to those of the rigid modules [Fig. 7(c)].

**Fig 7.**
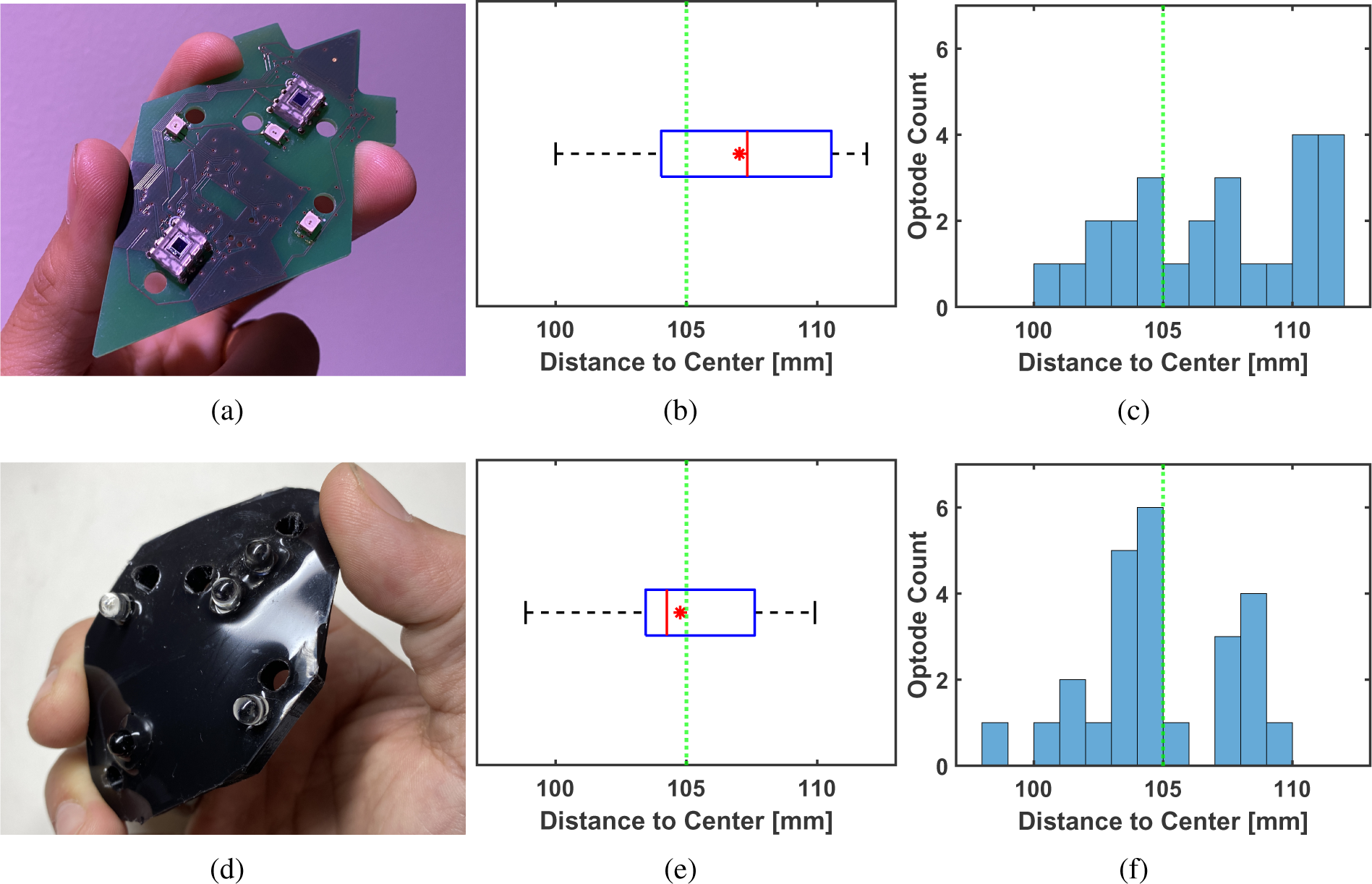
Comparison between (a) a rigid and (d) flexible module implementation in terms of conformity to a hemisphere phantom. Specifically, we report the distances to the sphere center measured for the (b) rigid and (e) flexible 5-module probes using a digitizer. The histogram of the resulting distances for a total of 25 optodes are shown for the (c) rigid and (f) flexible probes. With a 5 mm module thickness, a reading of 105 mm is expected for a perfectly conforming probe.

### 3.4 Evaluating the accuracy of automatic optode 3-D positioning

In Fig. 8(c), we plot and compare the 3-D optode positions acquired using the Polhemus digitizer against those computed using the MOBI module’s IMU readings and a piece-wise spherical model. The IMU-derived sources (red cross) and detector (blue cross) positions are overlaid on the manually digitized positions (cyan circles). Fig 8(d) plots the error between manually and automatically determined optode positions across all 25 optodes of the probe, indicating an average error of 4.2 mm, and Fig 8(d) shows the breakdown by module. In comparison, the standard deviation across the five repetitions of digitizer readings is 1.8 mm.

**Fig 8.**
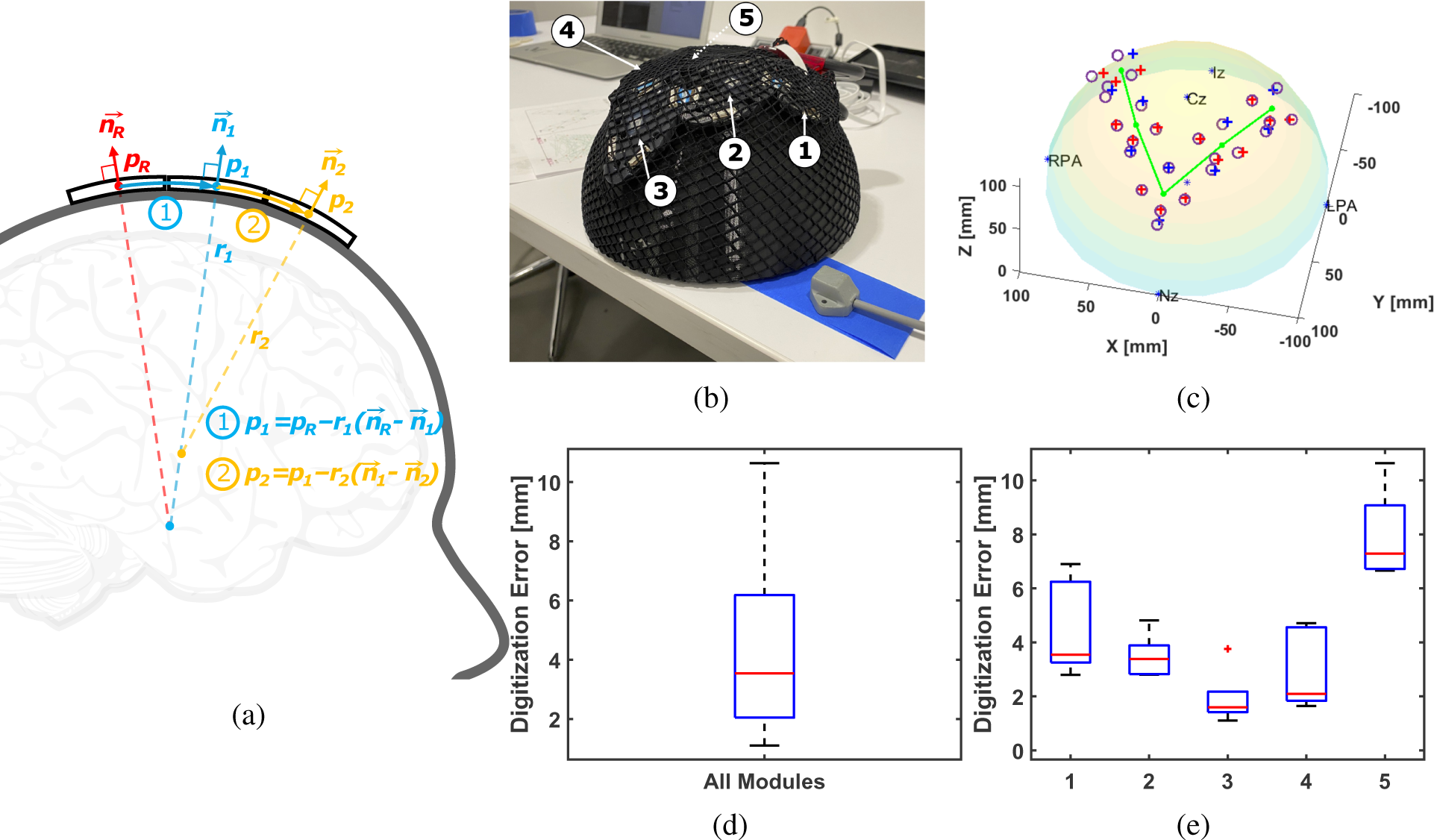
Schematic and results for automated optode position estimation, showing (a) a diagram of the piecewise-spherical algorithm to sequentially estimate module positions, p*_i_* (i = 1, 2, …), from a reference module location, p*_R_*, using their quaternion-derived unitary normal vectors, 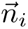, and the computed radius r*_i_* between adjacent modules; (b) a photo of a V-shaped MOBI probe secured by a hair-net over a hemisphere phantom; (c) estimated module positions (green) and optode positions (red - source, blue - detector) using IMU data versus manually digitized positions (purple circles). We also report the error of IMU-derived optode positions (d) on the probe level and (e) per module.

### 3.5 Physiological measurements of venous and arterial occlusion

We plot the estimated changes in hemoglobin concentrations during the dual-pressure blood cuff occlusion experiment in Fig. 9. During venous occlusion, oxygenated (*HbO*) and deoxygenated (*HbR*) hemoglobin show increasing trends in the results from both systems [Figs. 9(a) and 9(b)]. The arterial occlusion resulted in a negative correlation between HbO and HbR, as demonstrated by the horizontal total hemoglobin concentration (*HbT*) line. Upon releasing pressure, there is a brief increase in *HbO* and *HbT* accompanied by decreasing *HbR*.

**Fig 9.**
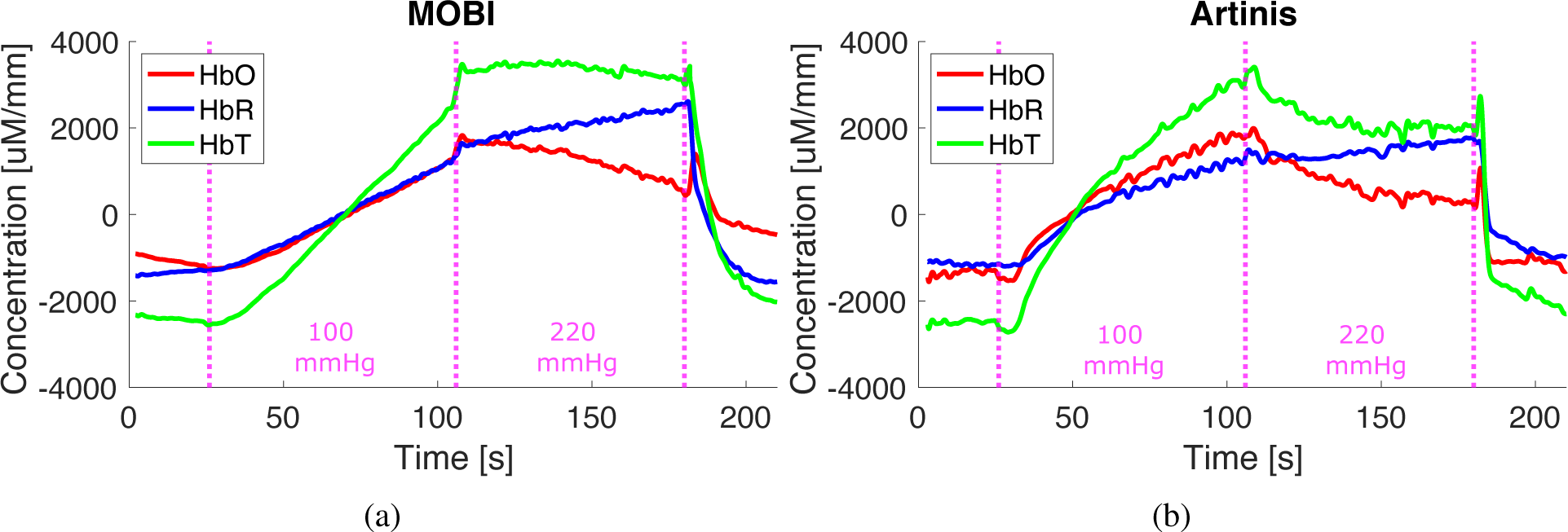
Results from a dual-pressure blood occlusion experiment using (a) a single MOBI module and (b) a single Artinis channel placed on the forearm. Venous (100 mmHg) and arterial (220 mmHg) occlusions lasted 75 seconds each prior to release.

### 3.6 Evoked response in motor execution task

In Fig. 10(a), we include a photo showing two modules affixed to the MOBI 3-D printed head-cap for the finger-tapping experiment while Fig. 10(b) provides the HRFs corresponding with each channel. A notable hemodynamic response is shown between detector D1 and source S2 in the contralateral (right) hemisphere, with a peak increase in *HbO* of 40 µM mm approximately 8 seconds after stimulus onset.

**Fig 10.**
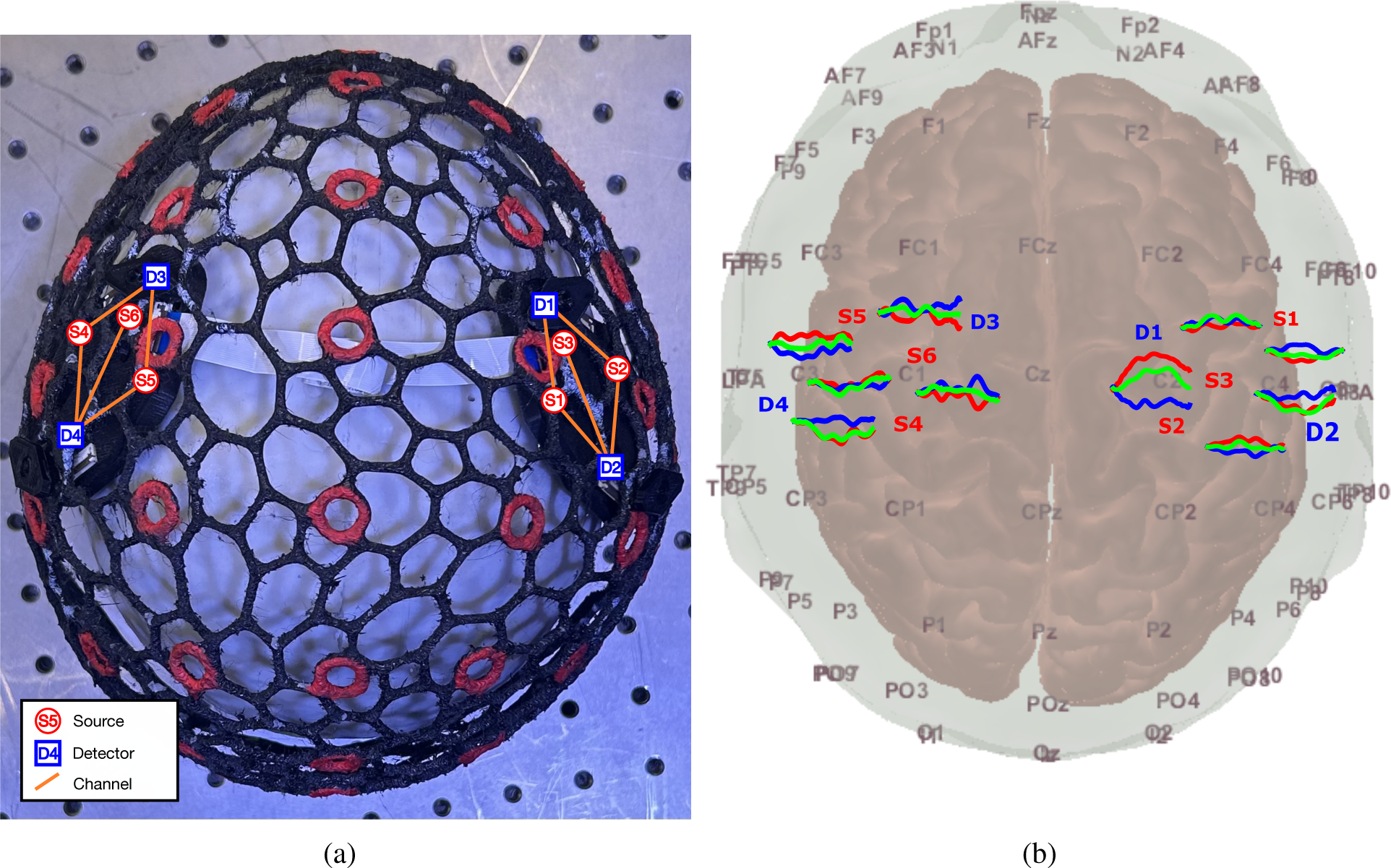
Validation of MOBI system in a finger-tapping experiment, showing (a) a photo of a two-module probe mounted over the wire-frame head-cap and (b) resulting hemodynamic responses for each channel, with red, green, and blue lines indicate HbO, HbR, and HbT changes, respectively. Source LEDs (red) and detectors (blue) are numbered in both plots.

## 4 Discussion

While modular fNIRS devices convey critical advantages in portability, wearability, and re-configurability, the MOBI system presented here is, to the best of our knowledge, the first 3-D aware and fully fPCB-based system. These features are expected to further facilitate the translation of fNIRS from laboratory and research settings to real-world applications and provide an expanded toolkit that empowers fNIRS studies in naturalistic environments. In combination with our 3-D printed, wire-frame head-cap, the flexible MOBI modules freely conform to any head shape, helping provide more robust optode-scalp contact even in environments with increased motion. Sampling from the IMUs embedded on each module also provides real-time, module-specific information of subject and probe movement while enabling the automatic determination of optode 3-D positioning. With further improvements, this 3-D optode localization function could readily enable tomographic fNIRS data analysis for more accurate localization of cerebrovascular activation while dramatically shortening probe setup time without the need for manual digitization. Additional support for wireless data collection over Wi-Fi and easy integration with data synchronization protocols like LSL uniquely position MOBI to aid the exploration of the human brain’s day-to-day functions.

Characterization of the MOBI system’s optical sensitivity, as shown in Fig. 5(a), reports an NEP of 7.3 and 8.9 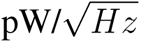 at 735 and 850 nm illumination, respectively, which is capable of measuring source-detector separations of up to 45-50 mm in human head tissue based upon previously reported studies.^40^ A dynamic range of *∼*88 dB, in conjunction with programmable source currents and detector gain settings that expand the effective range of measurable signals, also allows MOBI to simultaneously sample from long, inter-module channels as well as short separations channels for physiological noise regression^66^ without saturating the detectors.

The MOBI probe design pays specific attentions to enhanced portability, long-term wearability and accelerated experimental setup. The combination of the 3-D printable wire-frame head-cap, rapid clip-on module mounting, and automatic optode 3-D position sensing makes it possible to potentially shorten the experimental setup time by half or even more. With a measured 9.8 mΩ input resistance for each module, MOBI probes can be completely powered by a lightweight battery pack, theoretically capable of accommodating up to 75 modules on a 5 V power supply before dropping below the 3.3V necessary to drive the microcontrollers. Each module weighs only 18 g, meaning a 10 module probe, including head-cap, modules, master node, and a battery pack, would total less than 300 g, making MOBI well-suited for long-term wear.

Full-head modular probes not only require lightweight modules and large dynamic ranges but also an encoding strategy to ensure fast data frame rates. The results reported in Fig. 6 and Table 1 show how a spatial multiplexing encoding strategy can improve a full probe’s sampling rate. As long as sources are adequately spatially dispersed, their contributions to a detector’s readings avoid cross-talk. Table 1 also shows that tightly packed modules requires more SMGs, thus lowering the frame rate, compared to layouts that are more spatially separate even though the total number of modules remains constant. In this work, the SMG assignment is computed using a 2-D probe representation. It is also possible to perform this on a 3-D curved head-surface, although we anticipate a potential slight increase of required SMG numbers due to shortened distances. Although spatial multiplexing is a software feature and does not require extensive hardware, it does require knowledge of the location of all sources and detectors to identify the SMGs for a particular probe.

In addition, based on the comparisons between manual digitization shown in Fig. 7, it is not surprising that rigid PCB modules yield a higher average optode position than the expected value when compared to its flexible counterpart, as a result of poorer conformity to a curved surface. We want to note that previously reported rigid or flex-rigid modular fNIRS systems usually address such issues by using a smaller module size^36, 67^ or spring-loaded couplers^48^ in conjunction with flexible cables between rigid modules^41^ MOBI’s fully flexible modules offer broader intra-module source-detector separation ranges compared to modules with a smaller area and its optode-to-scalp coupling is expected to be further improved when combined with spring-loaded couplers similar to those used among other systems.

Fig. 8 provides initial validation for the accuracy of our internal IMU-based optode positioning system. It demonstrates that the average error for MOBI’s IMU-based optode position estimates is around 4.2 mm, and the calculation requires less than a second to complete. As expected, optode localization exhibits lower errors near the reference position (module 3) and increases when moving away from the reference. While this error is higher than that of repeated manual digitizations, which shows an average standard deviation of 1.8 mm over 5 repetitions, there are a few points of note. Firstly, inaccuracies in manually digitized optode positions may compound the error of the IMU-derived positions reported here. As shown Fig. 7(f), digitized positions for a flexible probe exhibits deviations of up to 5 mm from the expected distance of 105 mm between optodes and the center of the hemispherical phantom. The error of IMU-derived positions are computed with respect to each optode’s digitized positions, potentially resulting in an overestimation of said error. The higher localization error for Module#5 in Fig. 8(d) may be attributed to sub-optimal coupling that results from the non-uniform pressure applied by the elastic hair net when securing modules onto the large hemispherical dome phantom. The development of our 3-D printed headgear, as shown in Fig. 10(a) is specifically intended to address such issues in human studies. Finally, the piece-wise spherical approximation approach used to compute the position of each module with respect to other modules is a naive approach towards position estimation with respect to an unknown extrinsic reference using intrinsic quaternions. To further improve 3-D optode position estimation accuracy, we would consider strategies developed for the study of human kinemetics^68^ or optimization-based methods.^69^

We would like to note here as well that this method requires knowledge of the probe’s connection topology, inter-module distances, module geometry, and optode layout. While we use fixed FPC cable lengths to connect adjacent modules in this experiment, it is perceivable that experiments targeting spatially distant ROIs may require varying lengths of inter-module cables, which must be provided in order to obtain accurate 3-D optode positions. Additionally, anatomically-labeled headgear, such as our 3-D printed cap, serve as the bridge for registering the computed relative optode positions onto a head surface by allowing to affix the modular probe between desired head landmark positions. The use of emerging augmented reality and real-time head landmark tracking workflows could further shorten fNIRS probe setup time and improve probe placement accuracy.^70^ Although the use of orientation sensors in modular fNIRS systems has been previously reported for motion artifact correction,^71–73^ our reported results are among the first for extending IMU readings for 3-D probe shape acquisition.

Preliminary experiments in human subjects demonstrate that the MOBI system is capable of recovering changes in human physiology. Fig. 9 shows that simultaneous measurements using MOBI and a commercial fNIRS system during an arm cuff occlusion experiment recover similar hemodynamic changes compared to previous research results,^39^ such as the continuous increase of *HbO* and *HbT* during venous occlusion at 100 mmHg and a flattened *HbT* during arterial occlusion at 220 mmHg. In addition, both systems capture the transient increase in *HbO* following the release of arterial occlusion. Results from a single subject during a finger tapping task also produce a distinctive and expected hemodynamic response^74^ in the contralateral hemisphere following physiological signal regression using GLM.

Future work on the MOBI platform includes applying MOBI in experiments with more complex tasks, particularly in settings involving greater subject mobility. This would also allow further evaluation of the accuracy of IMU-derived versus manually digitized optode positions and assess MOBI’s ability to capture changes in probe/optode movement over the course of an experiment. In addition, the data acquisition frame rate is currently limited by the relatively low sampling rate and bit-depth of the ADC (12-bit at 32 ksps) included in the microcontroller. This limitation can be addressed through adopting higher performance sources and detectors. Increasing the source LED power, while still keeping it under safety limits, and adopting a high speed ADC in the detection hardware would lead to significant improvements in data frame rate. We are also considering using an analog front-end (AFE) to enable signal amplification, improve sampling rates, and perform on-board signal filtering. Any resulting improvements to SNR and sensitivity would also enable measurements across longer source-detector separations and thereby increase sampling density.

## 5 Conclusion

We have designed and validated a lightweight, fiber-less, diamond-shaped modular fNIRS system with features tailored toward use in natural environments. Its lightweight and flexible-circuit-based module design allow the system to conform to the scalp for improved optode-scalp coupling during use. In addition, a peer-to-peer communication network allows for the automatic determination of modular connection topology for the implementation of spatial multiplexing groupings to increase the frame rate of any probe configuration relative to sequential encoding methods. This, in conjunction with the 3-D orientation sensors embedded on each module, also allows for the determination of optode positions without the need for external hardware. Our MOBI modules were validated against a commercial system in blood pressure-cuff occlusion and finger-tapping tests. The MOBI fNIRS system directly addresses ergonomic considerations, enables fast experimental setup, and achieves the robust measurements necessary for advancing fNIRS towards further tackling challenges in naturalistic neuromonitoring tasks.

## Data Availability

All data produced in the present study are available upon reasonable request to the authors.

## Code, Data, and Materials Availability

Our open-source MATLAB toolbox for designing and optimizing modular fNIRS probes,^43^ as used in this work, can be freely accessed at http://github.com/COTILab/MOCA.

## Acknowledgments

The authors would like to thank Dr. David Boas for insightful comments and recommendations regarding improving signal quality. This research is supported by the National Institutes of Health (NIH) grants R01-EB026998 and R01-GM114365.

Biographies and photographs of the other authors are not available.

